# Spatially-resolved tumour infiltrating immune cells and prognosis in breast cancer

**DOI:** 10.1101/2024.07.22.24310819

**Authors:** Aaron J. Bernstein, Renske Keeman, Amber Hurson, Fiona M. Blows, Manjeet K. Bolla, Jodi L. Miller, Roger Milne, Hugo Horlings, Alexandra J. van den Broek, Clara Bodelon, James Hodge, Alpa Patel, Lauren R. Teras, Federico Canzian, Rudolf Kaaks, Hermann Brenner, Ben Schöttker, Sabine Behrens, Jenny Chang-Claude, Tabea Maurer, Nadia Obi, Fergus Couch, H. Raza Ali, Carlos Caldas, Irene Andrulis, Gord Glendon, Anna Marie Mulligan, Wilma Mesker, Agnes Jager, Annette Heemskerk-Gerritsen, Peter Devilee, Scott M. Lawrence, Jolanta Lissowska, Karun Mutreja, Thomas Ahearn, Stephen Chanock, Maire A. Duggan, Diana Eccles, J. Louise Jones, Will Tapper, Antoinette Hollestelle, Maartje Hooning, John Martens, Carolien H.M. van Deurzen, Angela Cox, Simon S. Cross, Mikael Hartman, Jingmei Li, Thomas C. Putti, Ute Hamann, Muhammad Rashid, Ania Jakubowska, Nicki Camp, Melissa H. Cessna, Amy Berrington de Gonzalez, Katarzyna Bialkowska, Jacek Gronwald, Jan Lubiński, Siddhartha Yadav, Pietro Lio, Doug F. Easton, Mustapha Abubakar, Montse Garcia-Closas, Paul D.P. Pharoah, Marjanka K. Schmidt

## Abstract

**Background:** The immune response in breast tumors has an important role in prognosis, but the role of spatial localization of immune cells and of interaction between subtypes is not well-characterized. We evaluated the association between spatially-resolved tissue infiltrating immune cells (TIICs) and breast cancer-specific survival (BCSS) in a large multi-center study.

**Patients and methods:** Tissue micro-arrays with tumor cores from 17,265 breast cancer patients of European descent were stained for CD8, FOXP3, CD20, and CD163. We developed a machine learning-based tissue-segmentation and immune cell detection algorithm using Halo™ to score each image for the percentage of marker-positive cells by compartment (overall, stroma, or tumor). We assessed the association between log transformed TIIC scores and BCSS using Cox regression.

**Results:** Total CD8+ and CD20+ TIICs (stromal and intra-tumoral) were associated with better BCSS in women with ER-negative (HR per standard deviation = 0.91 [95% CI 0.85 – 0.98] and 0.89 [0.84 – 0.94] respectively) and ER-positive disease (HR = 0.92 [95% CI 0.87 – 0.98] and 0.93 [0.86 – 0.99] respectively) in multi-marker models. In contrast, CD163+ macrophages were associated with better BCSS in ER-negative disease (0.94 [0.87 – 1.00]) and a poorer BCSS in ER-positive disease 1.04 [0.99 – 1.10]. There was no association between FOXP3 and BCSS. The observed associations tended to be stronger for intra-tumoral than stromal compartments for all markers. However, the TIIC markers account for only 7.6 percent of the variation in BCSS explained by the multi-marker fully-adjusted model for ER-negative cases and 3.0 percent for ER-positive cases.

**Conclusions:** The presence of intra-tumoral and stromal TIICs is associated with better BCSS in both ER-negative and ER-positive breast cancer. This may have implications for the use of immunotherapy. However, the addition of TIICs to existing prognostic models would only result in a small improvement in model performance.

**Highlights:** Stromal and intra-tumoral CD8+ and CD20+ TIICs are associated with better survival in ER+ and ER-breast cancers.

Stromal and intra-tumoral CD163+ TIICs are associated with better survival in ER- and poorer survival in ER+ breast cancers.

The presence of FOXP3+ tissue infiltrating lymphocytes in breast tumors was not associated with survival in breast cancer.

## Introduction

Survival after a diagnosis of breast cancer is affected by many patient and tumor characteristics, including age at diagnosis, tumor size, tumor grade, local and regional lymph node status, and expression of tumor biomarkers including estrogen receptor (ER), progesterone receptor (PR), KI67 and HER2. Several multiparameter molecular tests have been developed to aid prognostication and treatment decision-making in breast cancer [1]. Beyond the molecular characteristics of the neoplastic parenchyma, there is accumulating evidence that non-tumor cells might also influence disease prognosis.

A wide variety of tissue infiltrating immune cells (TIICs), such as cytotoxic T-lymphocytes (CD8+), T-helper lymphocytes (CD4+), B-lymphocytes (CD20+), natural killer cells (NK cells), and macrophages [2, 3], characterize the immune landscape of tumors. TIICs can occur in direct, cell-to-cell contact with tumor cells (intra-tumoral TIICs) or within the connective tissue stroma surrounding tumor cells (i.e., stromal TIICs) [4, 5]. Although the prognostic associations of TIICs have been evaluated across several studies, these were limited by a wide range of study designs with varying endpoints, sample sizes, metrics for immune cell infiltration, and statistical methodologies. As a result, the findings of these studies are inconsistent, but some broad patterns have emerged. Total tissue infiltrating lymphocytes are associated with higher pathological complete response to neo-adjuvant chemotherapy and improved disease-free survival in women with triple-negative breast cancer (TNBC) or HER2-positive breast cancer [5-7]. Of the specific immune cell types, CD8+ T-lymphocytes have been studied the most. A 2023 meta-analysis based on 14 studies found that CD8+ TIICs were associated with improved overall survival and disease-free survival, with similar associations for both intra-tumoral and peri-tumoral CD8+ TIICs [8]. They have also been found to be associated with better outcomes for women with ER-negative tumors (both triple-negative and HER2-positive) [9], ER-positive/HER2-negative tumors [9], and ER-positive tumors overall [10]. FOXP3+ T-lymphocytes have been associated with improved pathological complete response to neoadjuvant chemotherapy and overall survival in triple-negative and HER2-positive breast cancer, with some evidence that this effect is restricted to FOXP3+ TIICs in the stromal compartment [11]. Tissue infiltrating B-lymphocytes (CD20+) have been associated with better outcomes with little data on either ER-status specific effects or on tissue compartment effects [12]. One study reported that tissue infiltrating B-lymphocytes are associated with better outcomes for both HER2-positive and triple-negative breast cancer [13]. In contrast, tissue infiltrating macrophages (CD68+ or CD163+) have been found to be associated with worse overall survival and progression-free survival [14].

A few studies have evaluated more than one immune cell type in breast cancer. A high ratio of CD8+: to FOXP3+ TIICs has been associated with better disease-free survival in triple-negative [15, 16] and HER2-positive [15] breast cancers, and worse disease-free survival in luminal (ER-positive) breast cancers [17]. One study used deconvoluted bulk gene expression data to evaluate the role of all the above markers (CD8, FOXP3, CD20, CD163, and CD68) [18]. Increased CD20+ was associated with improved outcomes in ER-positive breast cancer with a similar association for CD20+ and CD8+ lymphocytes in ER-negative breast cancers. Poorer outcomes were observed for FOXP3+ lymphocytes and CD68 macrophages in ER-positive disease, and FOXP3+ lymphocytes and CD163+ macrophages in ER-negative disease.

The aim of this study was to clarify the prognostic associations of subsets of TIICs, specifically CD8+, FOXP3+, CD20+ and CD163+, in ER-positive and ER-negative breast cancer by using machine learning algorithms for the high-throughput, quantitative, assessment of TIICs, including their spatial localization within tissues. We addressed these aims in a large, multi-center study comprising over 17,000 patients with clinically annotated breast cancer tissues on TMAs that were stained using IHC for four markers representing the major immune cell subtypes.

## Methods

### Patient samples

Twenty-two studies from the Breast Cancer Association Consortium (bcac.ccge.medschl.cam.ac.uk) provided 323 tissue micro-arrays for staining for CD8, FOXP3, CD20, and CD163 as part of a larger project (B-CAST) to generate molecular pathology data on 15 markers in approximately 20,000 breast tumors.

Arrayed tumors were from female breast cancer cases from Canada, Germany, the Netherlands, Poland, Singapore, the UK, and the USA diagnosed between 1961 and 2015 (Table 1). Clinico-pathological data were available for ER status, age at diagnosis, tumor size, number of positive lymph nodes, vital status, cause of death, and follow-up time; methods of ascertaining vital status and cause of death are summarized in Supplementary Table 1. Cases diagnosed before 1980, missing vital status or age at diagnosis, or without any follow-up time after entry into the study were excluded, resulting in a total of 17,265 cases in the analyses. Of these a further 1,551 were missing ER status.

**Table 1:** Summary of contributing studies.

| Study | Country | Cases (N)* | Year of diagnosis* | Age at diagnosis | Tissue micro-arrays |  |
| --- | --- | --- | --- | --- | --- | --- |
|  |  |  |  | Mean [Min, Max] | Core size (mm) | Arrays (N) |
| ABCS | Netherlands | 461 | 2003- 2011 | 42 [18, 49] | 0.6 | 14 |
| UKBGS | UK | 629 | 2004-2014 | 56 [24, 84] | 1 | 28 |
| CPS2 | USA | 420 | 1993-2009 | 71 [51, 87] | 1 | 7 |
| EPIC | Germany | 263 | 1992-2000 | 58 [40, 75] | 1 | 9 |
| ESTHER | Germany | 258 | 2001-2004 | 62 [50, 75] | 1 | 6 |
| GESBC | Germany | 265 | 1992-1995 | 43 [20, 50] | 1 | 5 |
| MARIE | Germany | 1,484 | 2001-2005 | 62 [49, 75] | 1 | 30 |
| MCBCS | USA | 196 | 2001-2008 | 56 [26, 83] | 0.6 | 4 |
| MMH | USA | 151 | 2003-2013 | 66 [45, 89] | 0.6 | 4 |
| NEAT | UK | 1,944 | 1996-2001 | 49 [26, 76] | 0.6 | 16 |
| OFBCR | Canada | 199 | 1986-2002 | 48 [26, 78] | 0.6 | 3 |
| ORIGO | Netherlands | 483 | 1996-2006 | 53 [22, 86] | 0.6 | 11 |
| PBCS | Poland | 1,275 | 1998-2003 | 56 [27, 75] | 1 | 22 |
| PLCO | USA | 697 | 1995-2010 | 68 [55, 85] | 1 | 24 |
| POSH | UK | 835 | 2000-2007 | 36 [19, 41] | 1 | 39 |
| RBCS | Netherlands | 616 | 1980-2009 | 44 [25, 84] | 0.6 | 15 |
| SBCS | UK | 108 | 2012-2015 | 59 [24, 87] | 0.6 | 4 |
| SEARCH | UK | 4,593 | 1990-2009 | 53 [23, 73] | 0.6 | 32 |
| SGBCC | Singapore | 178 | 2007-2013 | 54 [25, 81] | 1 | 2 |
| SKKDKFZS | Germany | 816 | 1993-2005 | 60 [27, 93] | 0.6 | 19 |
| SZBCS | Poland | 1787 | 2002-2010 | 55 [31, 85] | 0.6 | 4 |
| UBC | USA | 1,236 | 1980-2009 | 60 [25, 93] | 2 | 25 |
| All |  | 17,265 | 1980-2015 | 54 [18, 93] |  | 323 |

\* After exclusion of cases diagnosed before 1980, missing vital status or age at diagnosis, or without any follow-up time after entry into the study

### Immunohistochemistry

Tissue microarrays are paraffin blocks in cassettes containing multiple cylindrical cores of tumor with a diameter between 0.6mm and 2mm, extracted from formalin-fixed and paraffin-embedded surgical tumor tissue blocks. The specific locations from which the cores were taken were determined by pathologists for each study to be the most representative of the overall tumor structure/composition. Eleven studies used 0.6 mm cores, ten studies used 1 mm cores and one study used 2 mm cores. Tissue microarrays were sent to the B-CAST co-ordinating centre at the University of Cambridge for sectioning and staining by the Histology Core at the Cancer Research UK Cambridge Institute. CD8, FOXP3, CD20, and CD163+ were selected as markers of key components of the immune response: CD8+ for cytotoxic response/immune upregulation, FOXP3+ for cytotoxic response/immune downregulation, CD20+ for humoral immunity, and CD163+ for innate immunity/immune downregulation. Details of the antibodies used for each marker are presented in Supplementary Table 2. The stained sections were then scanned at 20X using a Leica Aperio AT2 scanner, and the images stored using the PathXL software. The scanned images for each stained tissue microarray section were de-arrayed using study-specific tissue microarray maps, and an image of each stained core was exported as *jpeg* file for downstream analysis. Tissue microarrays included more than one tissue core per case for about 50% of cases (Supplementary Table 3). The dataset comprised 128,308 stained cores (32,268 CD8; 32,060 FOXP3; 32,106 CD20; and 31,874 CD163) from 17,265 cases (Supplementary Table 4).

**Table 2:**
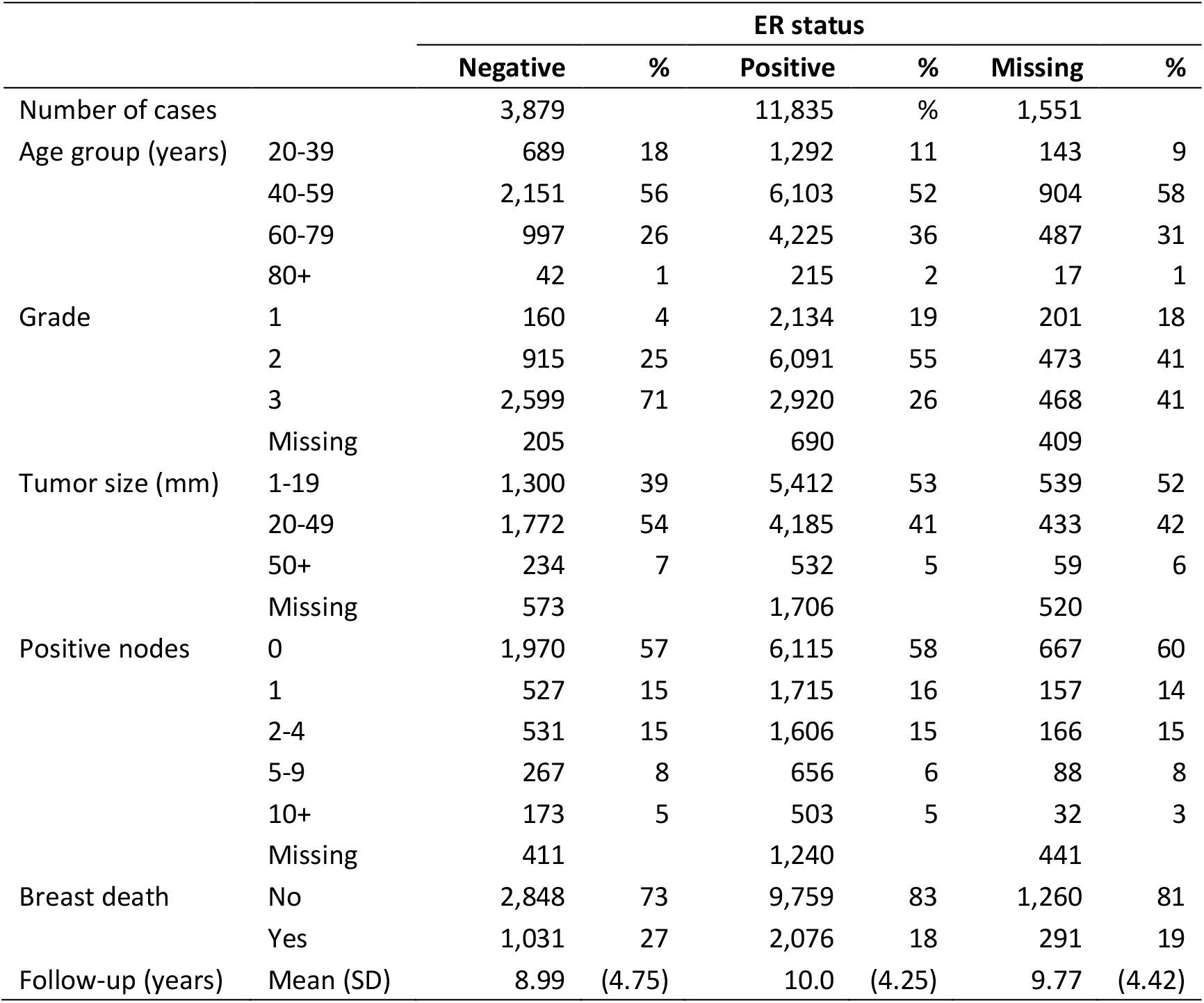
Characteristics of 17,265 cases by ER status.

### Image Analysis

Immune cell tissue infiltration scores were generated using Halo, a digital pathology platform produced by Indica Labs (Indica Labs, Albuquerque, NM). The image-analysis model was composed of two parts: an immune cell detection component and a tissue segmentation component (see Supplementary methods for details). Immune cell detection was performed by a proprietary immune cell detection script with minor hyperparameter optimisation. Development of the tissue segmentation component was an iterative process using a total of 553 core images (150 CD8+, 104 FOXP3+, 179 CD20+, 120 CD163+) annotated by pathologists (MAD, MA) to identify tumor, stroma, artefact, and glass (see Supplementary methods for details).

The automated algorithm was then applied to the complete set of images to generate the total area of tumor, stroma and artefact with the area of each occupied by TIICs. Individual core images were excluded from subsequent analyses if no tumor or stromal tissue was detected. The core-level immune cell score was then given as percentage of the compartmental tissue area occupied by TIICs. The number of cores per case varied between studies; case-level scores were taken as the mean value for cases with multiple valid tissue core scores. The automated scoring was validated against pathologist scores for CD8+ in the SEARCH study, as well as expert pathologist consensus on 80 validation images (20 per marker).

The maximum potential tissue area depends on the tumor core diameter and so the mean total tissue area detected by the tissue segmentation algorithm varied by tumor core diameter and by study (Supplementary figure 1). The proportion of cores with tissue area greater than 0.05 mm^2^, 0.1 mm^2^, 0.2 mm^2^ and 0.3 mm^2^ was 97 percent, 94 percent, 86 percent and 70 percent respectively.

**Figure 1:**
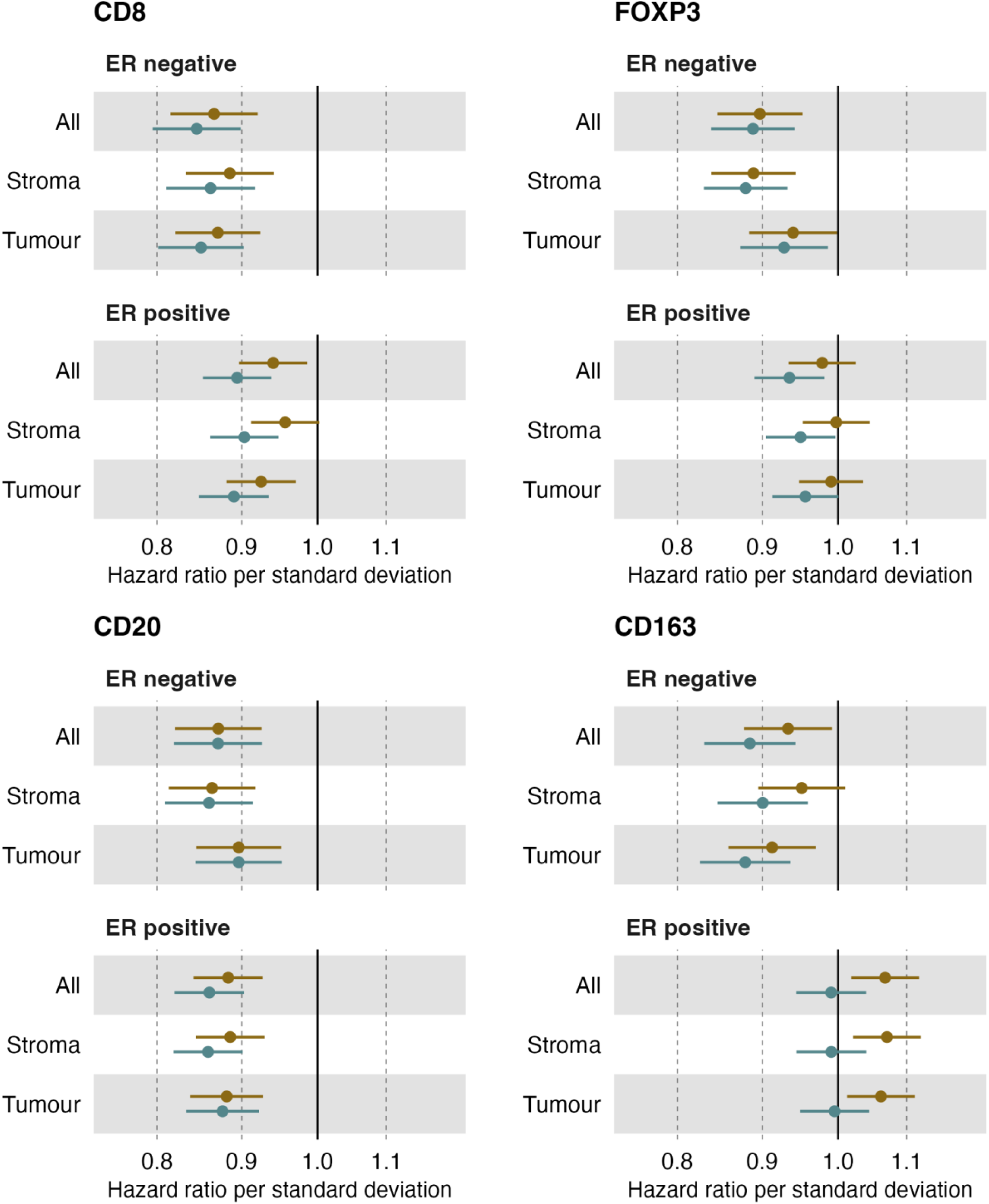
Hazard ratio for association between TIIC score and breast cancer specific survival by marker, ER status and tissue compartment in single marker models using imputed data. Brown = partially-adjusted model (adjusted for age and study), teal = fully-adjusted model (adjusted for age, tumor size, tumor grade, number of positive nodes and study).

### Statistical methods

Cox proportional hazards regression was used to assess the association between percent TIICs area and breast cancer specific survival (BCSS). Time at risk was from the date of diagnosis, with the time under observation beginning at the date at recruitment (left censoring). Follow-up was censored at death, the time of last observation, or 15 years after diagnosis, whichever came first. The event of interest was death from breast cancer. Where cause of death was not available (1,090 of 4,376 deaths), any death was treated as due to breast cancer. Separate analyses were performed for ER-positive and ER-negative tumors. Partially-adjusted models included age at diagnosis and study as covariates. Fully-adjusted models were adjusted for age at diagnosis, tumor grade, number of positive nodes, tumor size and study. Grade was found to violate the proportional hazards assumption (see Supplementary Methods) and so it was treated as a time-varying covariate with the log-hazard ratio varying as a function of log time. Participants from studies with fewer than 15 deaths were pooled into either a group of studies with 0.6 mm cores on the arrays or a group of studies with 1 mm cores on the arrays.

Age at diagnosis has been shown to have a non-monotonic hazard function for women with ER-positive breast cancers [19]. We, therefore, used the transformation of age at diagnosis described by Candido dos Reis and others for the ER-positive models which generates two variables for age [19]. The distribution of the TIIC scores (percentage of the tissue area occupied by TIICs) was highly skewed (Figure 2). We therefore log transformed the TIIC score and ran all the models twice using both untransformed and transformed scores. Partially-adjusted and fully-adjusted models were run for each marker individually and then multi-marker models including all four markers were run. TIIC scores based on a small area of tissue are likely to be unreliable, so we also evaluated the effect of using different thresholds for exclusion of cores based on area of tissue detected: we used thresholds for minimum tissue area of 0.05 mm^2^, 0.1 mm^2^, 0.15 mm^2^, 0.1 mm^2^, 0.25 mm^2^, and 0.3 mm^2^.

**Figure 2:**
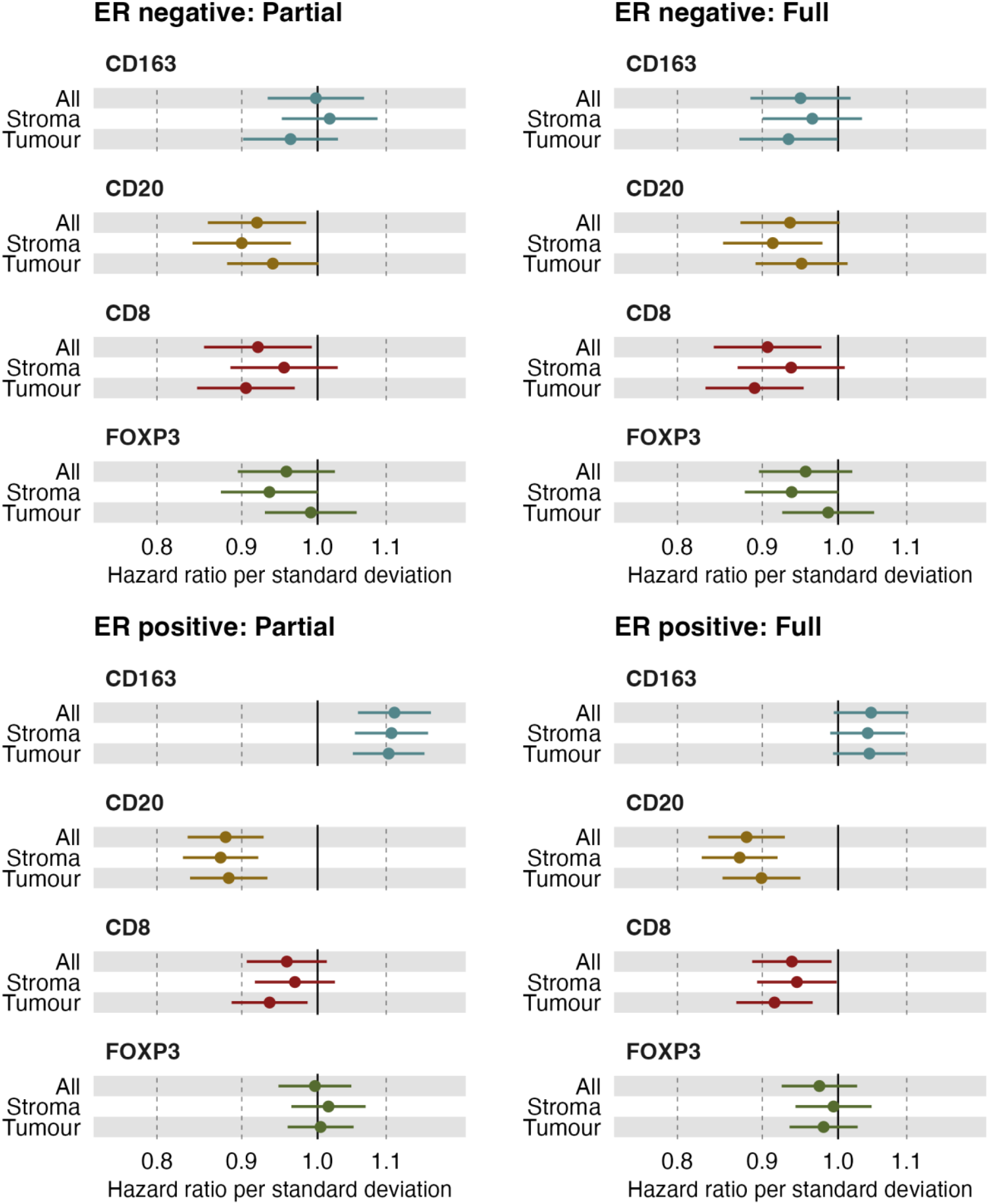
Hazard ratios for association between TIIC score and breast cancer specific survival by ER status, marker and tissue compartment for partially (adjusted for age and study) and fully-adjusted (adjusted for age, tumor size, tumor grade, number of positive nodes and study) multi-marker models using imputed datasets

Missing data for TIIC scores reduced the sample size of the complete case analysis (i.e. cases with missing data removed) for the partially-adjusted multi-marker model to 7,450 ER-positive cases and 2,539 ER-negative cases with the core exclusion threshold of 0.25 mm^2^. This was reduced further in the fully-adjusted models to 5,585 ER-positive cases and 1,960 ER-negative cases because of missing data for grade, tumor size and number of positive nodes. We therefore used multiple imputation by chained equations to impute missing data and maximise the sample size in all analyses. The scores for the four markers with study, age at diagnosis, ER status, grade, tumor size, number of positive nodes, follow-up time and breast cancer mortality were used in the imputation models, and each dataset was imputed 10 times. The results from the Cox regression analyses on each imputed data set were combined using Rubin’s rules [20].

We approximated the relative variation [21] accounted for by TIIC scores as

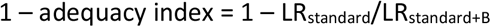

where LR is the likelihood ratio chi-squared statistic, the standard model includes age at diagnosis, ER status, grade, tumor size, number of positive nodes and B denotes the inclusion of TIIC scores as predictors.

We evaluated between study heterogeneity by running the fully-adjusted, multi-marker models for the imputed data using the 0.25 mm^2^ tissue area exclusion threshold separately for each study. We tested for evidence of between study heterogeneity using Cochran’s *Q* [22] and *I*^*2*^ as an estimate of the proportion of overall variance due to between study variance [23].

All analyses and data visualisations were carried out using R [24], implemented in R Studio [25], with the R packages *tidyverse* [26], *broom* [27], *ggforestplot* [28], *lemon* [29], *meta* [30], *mice* [31], *patchwork* [32], and *survival* [33].

## Results

There was a total of 128,308 core images from 17,265 patients after the application of initial case exclusion criteria. The patient and tumor characteristics of the cases are summarized in Table 2. The CD8+ TIIC scores were validated by comparing them to manual scoring by a pathologist (HRA) of 2,247 cores from the SEARCH study as previously reported [9]. A good correlation was observed for total score (Spearman’s Rho = 0.76), intra-tumoral score (Rho = 0.55) and stromal score (Kappa = 0.62) (Supplementary Figure 3).

The choice of untransformed or log-transformed TIIC scores was based on the results of an initial set of single marker, complete case analyses. We ran 576 single marker Cox regression models given all possible combinations of four markers, partially and fully-adjusted models, three tissue compartments, ER-positive and ER-negative models, six thresholds for minimum tissue area, and untransformed and transformed TIIC scores. There were 288 models for each of the analyses using the untransformed and log-transformed score variable. The log-transformed was the better model in 229, based on the model log likelihood. There were 96 models at each tissue area threshold with no single threshold being clearly the best fit with the 0.25 mm^2^ threshold being best in 24 models and the 0.05 mm^2^ threshold being best in 31 models. The results for all 576 complete case analysis models are provided in Supplementary Table 5.

Subsequent analyses were based on the data for all 17,265 cases after multiple imputation with the tissue area threshold on a minimum of 0.25 mm^2^ and the log transformed percent tissue area TIIC score. In single marker analyses, CD8+ TIICs in both tumoral and stromal compartments were associated with improved BCSS in ER-positive and ER-negative disease (Figure 1). The effect sizes were similar in the partially- and fully-adjusted models. FOXP3+ TIICs were associated with a better prognosis in ER-negative and in ER-positive cases, but the effect in ER-positive disease was only observed in the fully-adjusted models. CD20+ TIICs in both compartments were associated with better breast cancer specific survival in both ER-positive and ER-negative with no attenuation of the association in the fully-adjusted models. Finally, CD163+ TIICs in both compartments was associated with better prognosis in ER-negative cases. In ER-positive cases CD163+ TIICs were associated with a poorer prognosis in the partially-adjusted models, but the effect was completely attenuated after adjusting for grade, size and number of positive nodes. The results for the single marker fully-adjusted models did not vary substantially by tissue area exclusion threshold (Supplementary Figure 4 and Supplementary Table 5).

The results of the multi-marker, partially- and fully-adjusted models including all four immune cell markers using the imputed data are shown Figure 1: Hazard ratio for association between TIIC score and breast cancer specific survival by marker, ER status and tissue compartment in single marker models using imputed data. Brown = partially-adjusted model (adjusted for age and study), teal = fully-adjusted model (adjusted for age, tumor size, tumor grade, number of positive nodes and study).

Figure 2. CD8+ and CD20+ TIICs were associated with improved prognosis in both ER-positive and ER-negative disease with a slightly stronger effect for CD8+ infiltration in the tumor compartment. CD163+ TIICs were associated with a better prognosis in ER-negative cases and a poorer prognosis in ER-positive cases with similar effects in both stromal and tumoral compartments. FOXP3+ infiltration was no associated with outcome, although the confidence intervals were wide. Overall, there was some evidence of inter-study heterogeneity for CD20+ and CD163+ (**Error! Reference source not found**.), which might be expected given the underlying heterogeneity in the study designs including the year of diagnosis, the time of storage of pathology material and methods for TMA construction. The fraction of the variance explained by the model that was accounted for by the TIIC scores was 7.6 percent for the ER-negative model and 3.0 percent for the ER-positive model.

## Discussion

We evaluated the association with breast cancer-specific mortality of the presence of tissue infiltrating cytotoxic T-cells (CD8), regulatory T-cells (FOXP3), B-cells (CD20), and M2 macrophages (CD163), along with well-established prognostic factors in female breast cancer. No published studies have reported on the effects of all four of these markers in multi-marker models. CD8+ and CD20+ TIICs were associated with better survival in both ER-positive and ER-negative disease. CD163+ TIICs were associated with a better survival in ER-negative cases and a poorer survival in ER-positive cases. FOXP3+ lymphocyte infiltration was not associated with outcome, although the confidence intervals were wide. Our findings for the single marker analyses are broadly consistent with those in the literature, with some notable differences. We found better prognosis for CD8+ and CD20+TIICs in both ER-positive and ER-negative disease and for TIICs in both stromal and intra-tumoral compartments. Published studies have found that increased CD8+ TIICs predict better outcomes in ER-negative but not ER-positive breast cancer apart from the small subgroup of ER-positive that is also HER2-positive [8, 9]. While an association of CD20+ B-lymphocytes with survival for women with invasive breast cancer has been described previously, associations in either ER-negative or ER-positive disease have not been shown. An association of FOXP3+ TIICs with better survival is well established in ER-positive and ER-negative disease and is supported by our single marker analyses [11, 34]. We found CD163+ macrophage infiltration to be associated with a poorer survival in ER-positive patients although the association was completely attenuated on adjustment for other prognostic variables. This is similar to the few published studies that have reported on CD163+ macrophages in ER-positive disease [14]. In contrast, we found infiltrating CD163+ macrophages to be associated with an improved survival in ER-negative breast cancer, whereas most published studies of ER-negative or triple-negative breast cancers have reported a poorer prognosis for CD163+ TIICs [14]. The reasons for these differences are unclear. The difference between our results and those of previous studies for CD8+ and FOXP3+ in ER-positive cases is likely to be due to the substantially increased statistical power of our study to detect modest effects. The improved survival observed for CD163+ TIICs in ER-negative cases is harder to explain. Given the concordance of our findings for CD163+ TIICs with other studies in ER-positive cases it seems unlikely that bias can be an explanation. Chance is a possible but unlikely explanation, given the opposite direction of effect in our study.

The weaknesses of the study need to be considered when interpreting the findings. We assumed that 1,090 patients with an unknown cause of death – out of 4,376 deaths within 15 years of follow-up – died of breast cancer. This will be associated with some misclassification, particularly in the period 10-15 years after diagnosis when almost half of deaths are due to causes other than breast cancer (Supplementary Table 7). Consequently, this may result in under estimation of the effect sizes, though this is likely to be small, and the added power from the additional events will outweigh the effect of misclassification. Breast tumors are spatially heterogeneous, and the cores sampled for tissue microarrays cannot capture all of the heterogeneity. This is also likely to result in some under estimation of effect sizes. We explored the potential effect of this by comparing the results for two of the strongest single marker associations, that for CD8+ TIICs in both tissue compartments in ER-negative disease (HR_full_ = 0.83, P = 1.6 × 10^−6^) and CD20+ in tumor tissue of ER-positive disease (HR_full_ = 0.79, P = 1.8 × 10^−6^), in subsets of the data based on cases represented by a single core and cases represented by two or more core. The effect size for the CD8+ analysis was somewhat larger for the multi-core case sample (HR_full_ = 0.80, 95% CI 0.72 -0.89) than for the single core case sample (HR_full_ = 0.85, 95% CI 0.76 – 0.95). The difference between the samples for the CD20+ tumor infiltration score was more substantial (HR_full_ = 0.68 95% CI 0.58 – 0.81 compared to HR_full_ = 0.87 95% CI 0.87 – 0.98). This is consistent with the observation that B-lymphocytes tend to aggregate whereas T-cells do not [35].

Our study was based on samples of tumors of all sizes, grades and subtypes from several countries, with the majority of cases being of white European ancestries. The study is representative for the European population. Given the similar associations of the well-establish prognostic factors in diverse populations, it seems likely that the findings would also apply to other populations. We provided solid evidence to confirm known associations between CD8+, CD163+, and CD20+ TIICs and breast cancer survival in ER-negative disease and provided novel evidence for associations between those markers and ER-positive disease. This supports further consideration of inclusion of ER-positive patients in clinical trials of immune modulators.

## Data Availability

The tissue segmentation and TIIC scores generated by the Halo algorithm together with the phenotype data, the imputed datasets and the analysis code will be available at the European Genome Phenome Archive on publication (https://ega-archive.org/).

## Acknowledgements

The authors thank the Histology and Genomics Cores at the Cancer Research UK Cambridge Institute for technical support, and all patients, researchers, clinicians, and administrative personnel who contributed to the studies taking part in B-CAST. The CPS-II study investigators acknowledge the contribution to this study from central cancer registries supported through the Centers for Disease Control and Prevention’s National Program of Cancer Registries and cancer registries supported by the National Cancer Institute’s Surveillance Epidemiology and End Results Program.

BCAC was supported by Cancer Research UK grant: PPRPGM-Nov20\100002 and by core funding from the NIHR Cambridge Biomedical Research Centre (NIHR203312). The views expressed are those of the author(s) and not necessarily those of the NIHR or the Department of Health and Social Care. The B-CAST project was supported by the Horizon 2020 Research and Innovation Programs of the European Union B (grant number: 633784) and the NIHR Cambridge Biomedical Research Centre. AJB was supported by the NIH/Oxcam doctoral programme. The funding of the contributing studies is listed in Supplementary Table 8.

## Ethics

All participants provided written informed consent. The ethics committees or institutional review boards responsible for oversight of the individual studies are listed in Supplementary Table 9.

## Author contributions

Conceptualization and design: MG-C, PDPP, MKS

Study recruitment and sample collection: AA, HRA, IA, TA, AB, CB, HB, KB, SB, AC, CC, FCa, FCo, JC, MC, NJC, SSC, PD, DE, DFE, GG, JG, AH-G, MG-C, AHo, AHu, HH, MHa, MHo, AJag, AJak, JLJ, RK, JLi, JLis, JLu, SML, AMM, JM, JLM, KM, RM, TM, WM, NO, AP, TCP, PDPP, MR, BS, MKS, LRT, WT, AJvdB, CHMvD, SY

B-CAST co-ordination and data support: FMB, RK, MJB, RM

Immunostaining: JM

Pathology review: HRA, MA, MAD

Algorithm development: AJB, AMA, PL

Statistical analysis: AJB, PDPP

Interpretation: AJB, AMA, MG-C, PDPP, MKS

Manuscript writing of original draft: AJB, AMA, MG-C, PDPP, MKS

Final approval: all authors.

