## Supplementary methods and figures for "Spatially-resolved tumour infiltrating immune cells and prognosis in breast cancer"

### Supplementary Materials

#### 1 Supplementary figures referenced in main text

Supplementary figure 1: Boxplot and jitter plot of core tissue area (tumor plus stroma) by study and tumor core diameter

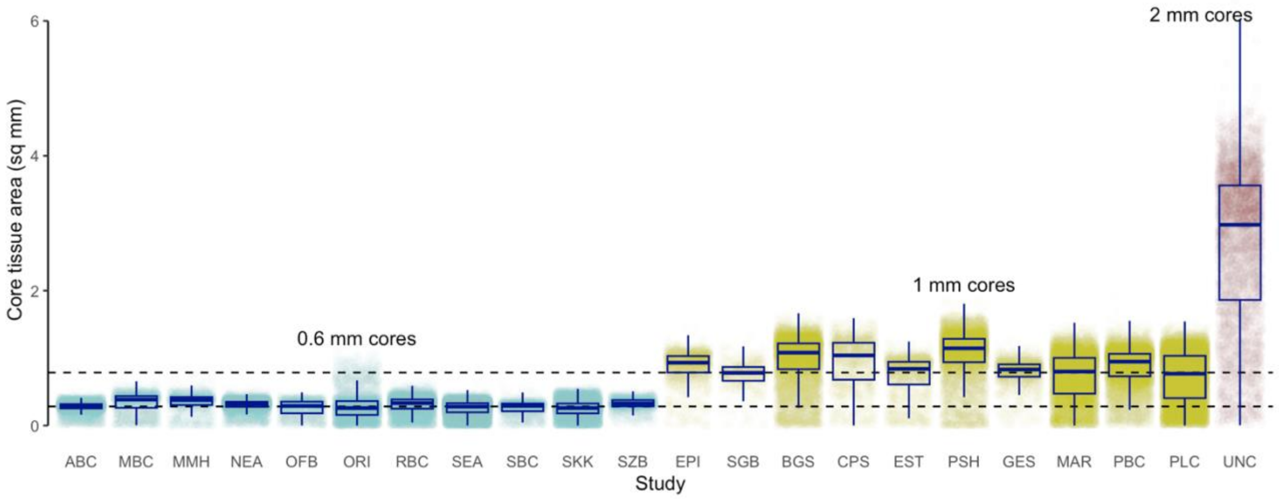


Supplementary figure 2: Boxplot of distribution of TIIC scores (percentage of the tissue area occupied by TIICs) by marker and tumor core size. Y-axis is log scale.

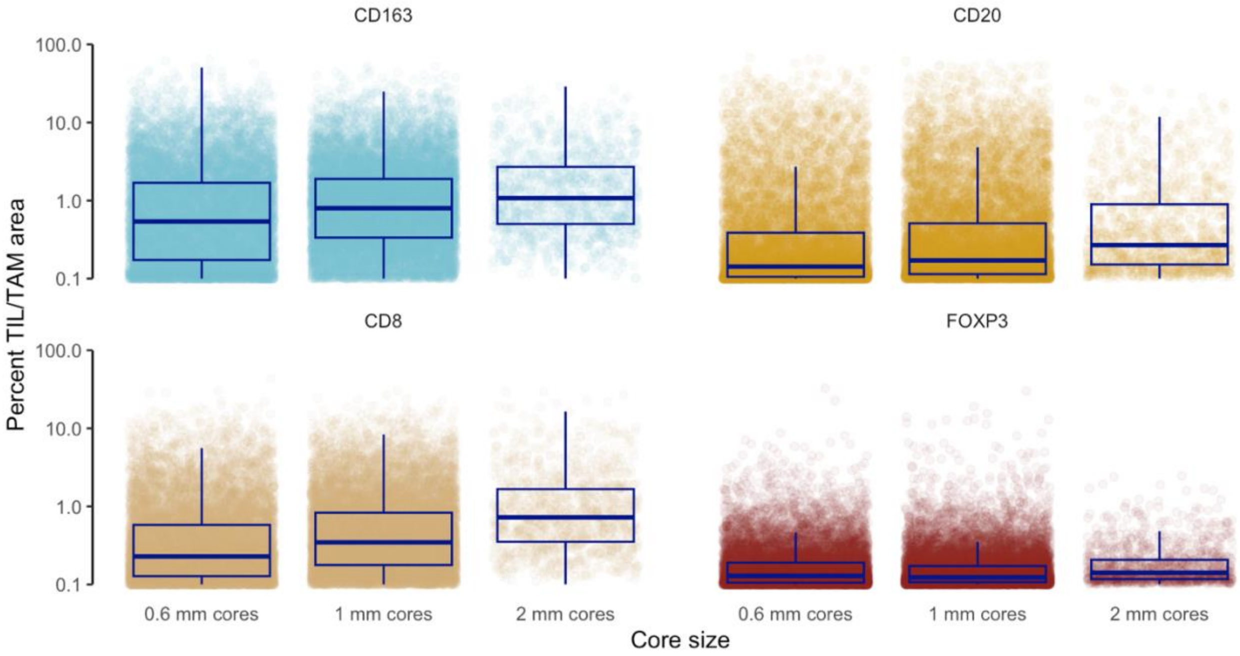

Supplementary figure 3: Scatterplots of pathologist CD8+ TIL scores v automated CD8+ TIL scores (percent tissue area) by tissue compartment

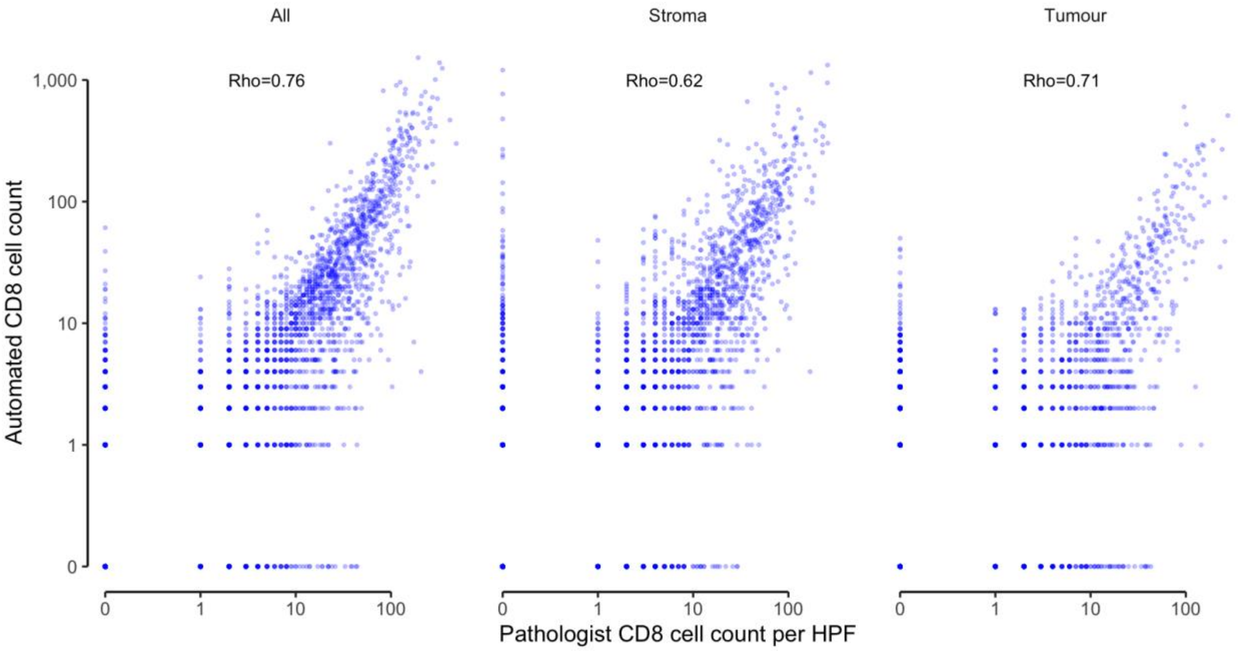

Supplementary figure 4: Hazard ratio for association between TIIC score and breast cancer specific survival by marker, ER status, tissue compartment and core exclusion threshold. Fully-adjusted single marker models with imputed data.

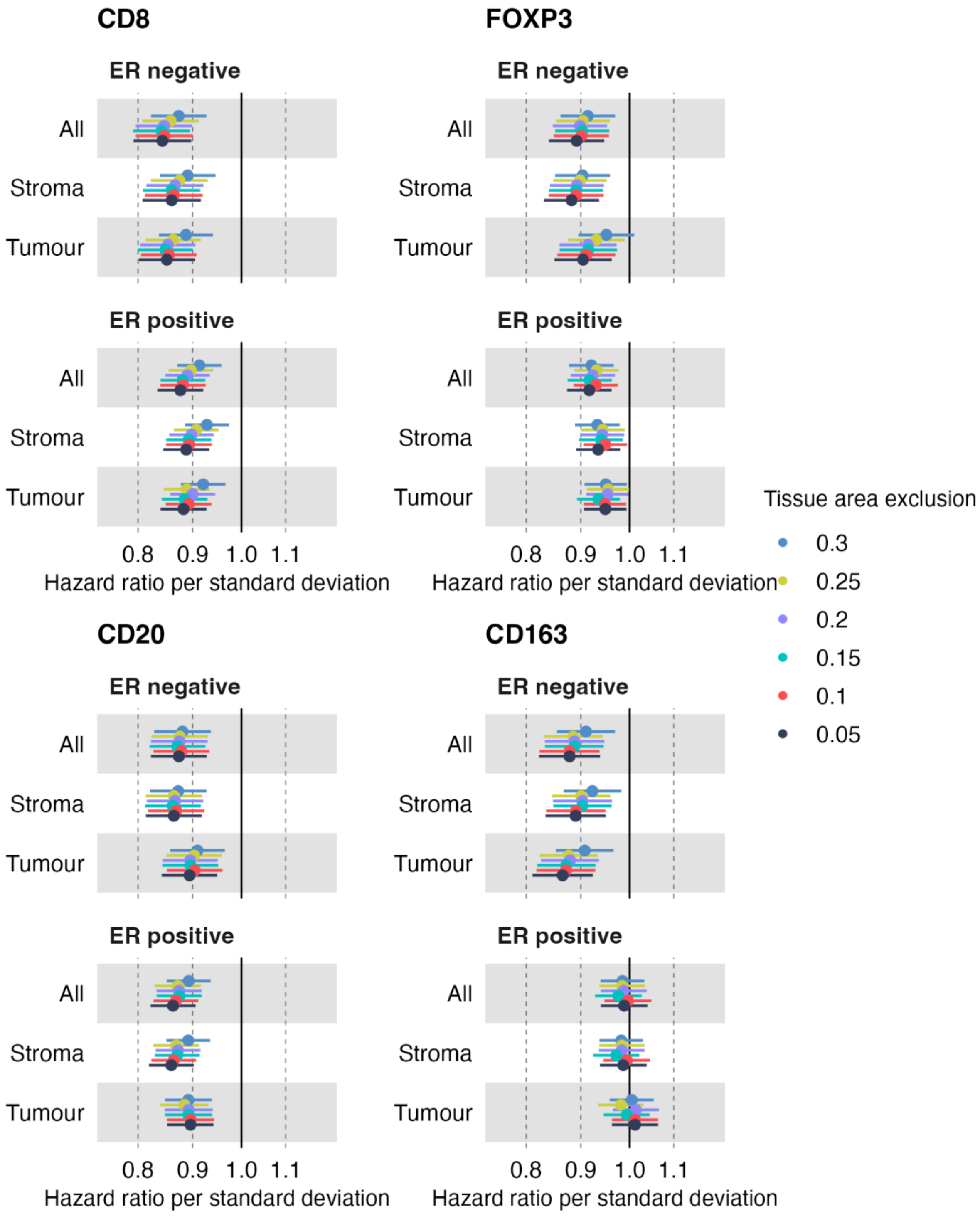

#### 2 TIL Detection Model Development

The primary functions of the model were first to segment the tissue into stromal and intratumoral compartments and second to detect TILs/Tams. The TIIC detection was done using a proprietary algorithm developed by Indica Labs. The model was used "off shelf" with minor manual tuning of hyperparameters to ensure a good visual fit on sample images. Hyperparameters were detection average RGB colour optical density (OD) for both the target stain (red OD = 0.268, green OD = 0.570, blue OD = 0.776) and the background stain (red OD = 0.644, green OD = 0.716, blue OD = 0.267); image zoom (0.9x); tissue edge thickness (0 $\mu m$); minimum tissue OD (0.037); minimum immune stain OD (0.13); membrane detection tolerance (0.13); and immune cell size range (5-270 $\mu m^2$).

#### 3 Tissue Segmentation Model Development

Halo uses MiniNet, a convolutional neural network (CNN), for tissue segmentation. However, both physical damage and digital artefacts were present in an unknown number of images. Physical damage includes darkly-stained debris (the most common artefact), tearing and folding of the thin core sections, and nonspecific antibody binding to non-target tissue during IHC staining (Supplementary figure 5A-B). Digital damage consisted of dearraying errors which result in adjacent cores being visible (and therefore scorable) within an image (Supplementary figure 5B). Thus, an additional requirement of the segmentation component of the model was to segment the various artefacts that would be subsequently excluded from the analysis. Hyperparameters for tissue and QC segmentation were minimum object size (140 $\mu m^2$) and resolution (1.15 $\mu m$ per pixel).

##### *3.1 Segmentation Model Training*

The segmentation model was initially trained on 14 annotated (manual segmentation) CD8 images from the BGS and SEARCH studies selected to include a mixture of both damaged and undamaged images with simple and complex phenotypes. This included images with lymphoid aggregates (Supplementary figure 5D) as the high TIL density in these phenotypes may result in misclassification as tumour (due to the absence of the collagen striations that typify stroma) or as artefact (due to the visual similarity between high-density TILs and darkly-stained debris). We also included examples of tumour-rich cores with an appearance that might be misclassified as stroma (Supplementary figure 5E).

The trained segmentation model was then run iteratively on 320 CD8 images selected to represent the diverse stain qualities, damage profiles, and histopathologic features that the model may encounter. The model was run on the first image; if there were any segmentation errors the image was re-annotated by hand (by AJB) to correct the errors, the image was added to the set of training images, and the model was then re-trained on the enlarged training set. This was repeated until all 320 images had been analysed. Twenty-nine of the 320 images required correction and were added to the model training set (14+29 = 43 images).

Supplementary figure 5: Example tumour core images

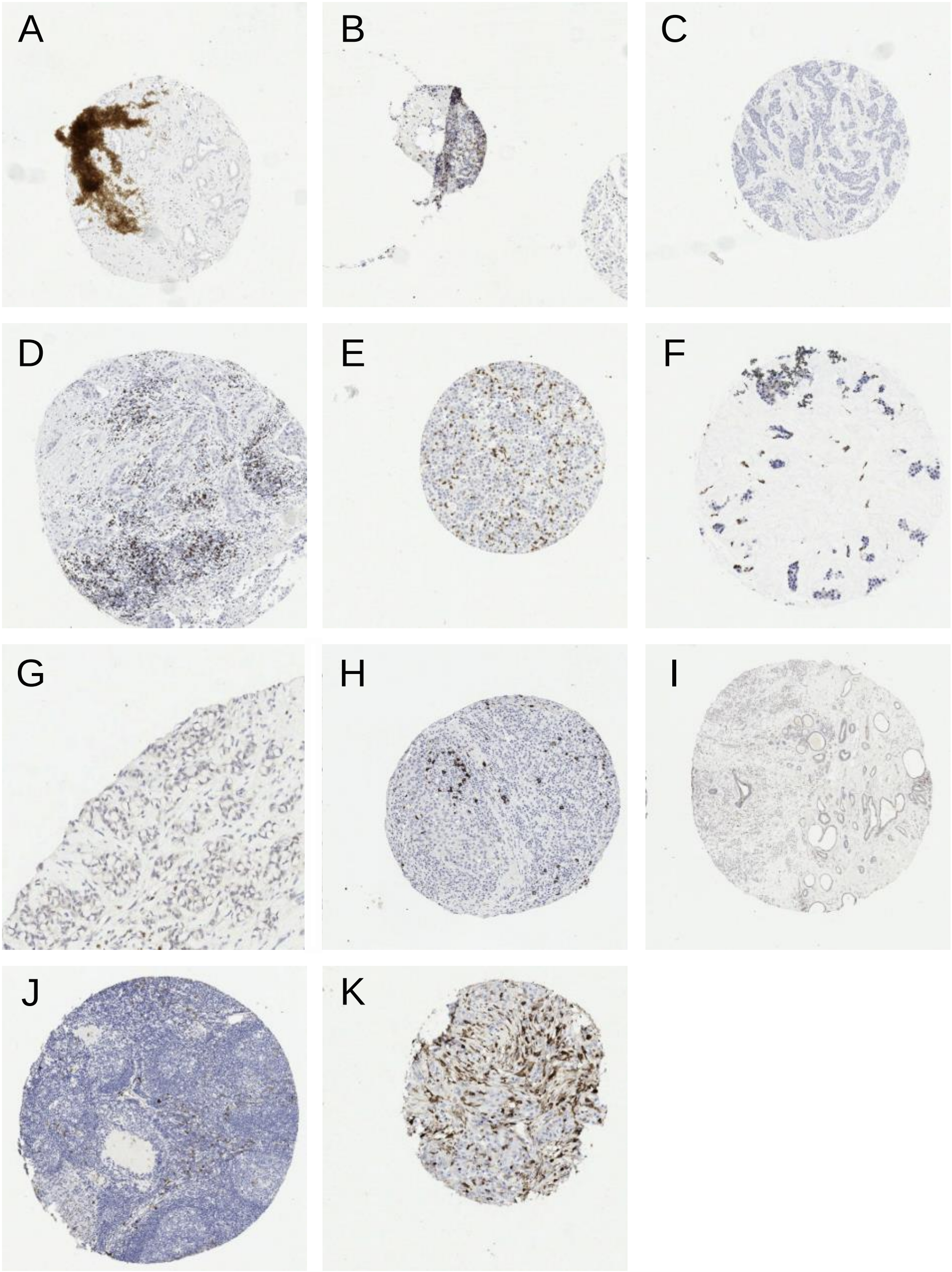

Next, we randomly sampled 1,800 images (approximately 20 images/study/marker) and followed the image-by-image process of running the model, re-annotating by AJB if required and adding to the training set before re-running the training. In total, 289 images were re-annotated (27 CD8, 60 FOXP3, 141 CD20 and 61 CD163) and added to the training set. The model was re-trained and the image-by-image analysis of another 1,800 random images was repeated from which 219 required re-annotation by AJB (79 CD8, 43 FOXP3, 38 CD20 and 59 CD163) before adding them to the training set

The training set then comprised 551 (14 + 29 + 289 + 219) images with segmentation annotations done manually by AJB. A random sample of 80 of these images (20/marker) and 101 images that had been identified during the first round of iterations to have features that made segmenting tumour and stroma inaccurate such as unusual pathologies (e.g. signet ring cell carcinoma or mucinous carcinoma [Supplementary Figure 1F-G]) or poor staining were then visually inspected by two pathologists (AMA and MAD). One hundred and forty-three of the annotations that had already been done manually by AJB were revised and another two of the problem images were annotated manually for the first time.

The final set of 553 images that had been hand segmented by either AJB (408 images) or the pathologists (145 images) was then split into training (442) and validation (111) sets in order to optimize the number of MiniNet iterations used for training based on the precision, recall and F1 statistics of the segmentation on the validation data. As the training iterations increase, the model would be expected to improve and then deteriorate as overfitting occurs. However, even after 200,000 iterations this was not observed and so we halted the training after ~32,000 iterations when it provided high-quality predictions on a few challenging images, enriched for tumour, rare breast cancer types (e.g., signet cell carcinoma), and lymphoid tissue [Supplementary Figure 1H-J].

The outputs of the model were area of tumour, stroma, and artefact output by the tissue segmentation component and TIIC area partitioned by tissue compartment output by the TIIC detection model.

##### *3.2 Tissue Segmentation Model Validation*

The final trained segmentation model together with the TIIC detection model was then run on a random set of 100 images from the training set (25 per marker). The automated tissue segmentation and TIIC detection were visually inspected by two pathologists (AMA and MAD) and assessed for quality. The quality of segmentation was based on a consensus rating of the severity of any tissue segment misclassification where overprediction of the tumour in the stroma was considered to be low severity and underprediction of true tumour was considered to be high severity. TIIC detection was considered successful if virtually all the TIICs were correctly identified (allowance for minor variance was made at the discretion of the participating pathologists). False positives were not considered, as that was the responsibility of the tissue segmentation's artefact. Ultimately, tissue segmentation was considered to pass in 85% of the images with TIIC detection passing in 98%.

Recall, precision, and F1 scores were then estimated. The algorithm was found to perform well, with average scores across all marker and compartment combinations of 0.91, 0.92, and 0.90, for recall, precision, and F1 scores, respectively.

###### *3.2.1 Quality Control and Sensitivity Analyses*

The segmentation algorithm was trained to distinguish artefact from stained tissue and the proportion of tissue on each core. The proportion of segmented tissue classified as artefact, at the patient level, ranged from 0 to 100 percent, with 4 percent having no artefact and 81% having less than 10 percent artefact. The amount of artefact on the CD163 stained images was slightly higher than that of the other three marker (Supplementary figure 5K). The percent tissue artefact correlated positively with TIIC infiltration (Supplementary figure 7) but there was little correlation between the tissue area and TIIC score

(Supplementary figure 8).

Supplementary figure 6: Jitter plot with superimposed boxplot of log(percent artefact+0.1) for each core by marker

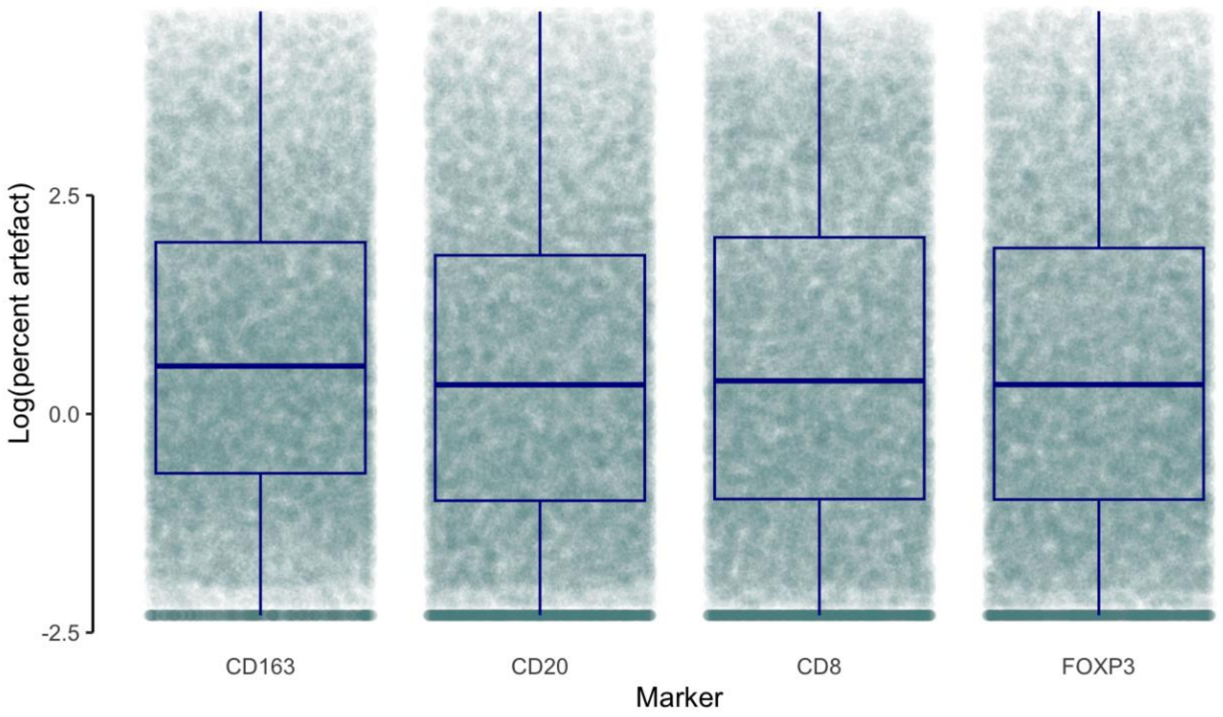


Supplementary figure 7: Scatterplot of percent tissue artefact against percent TIIC by marker

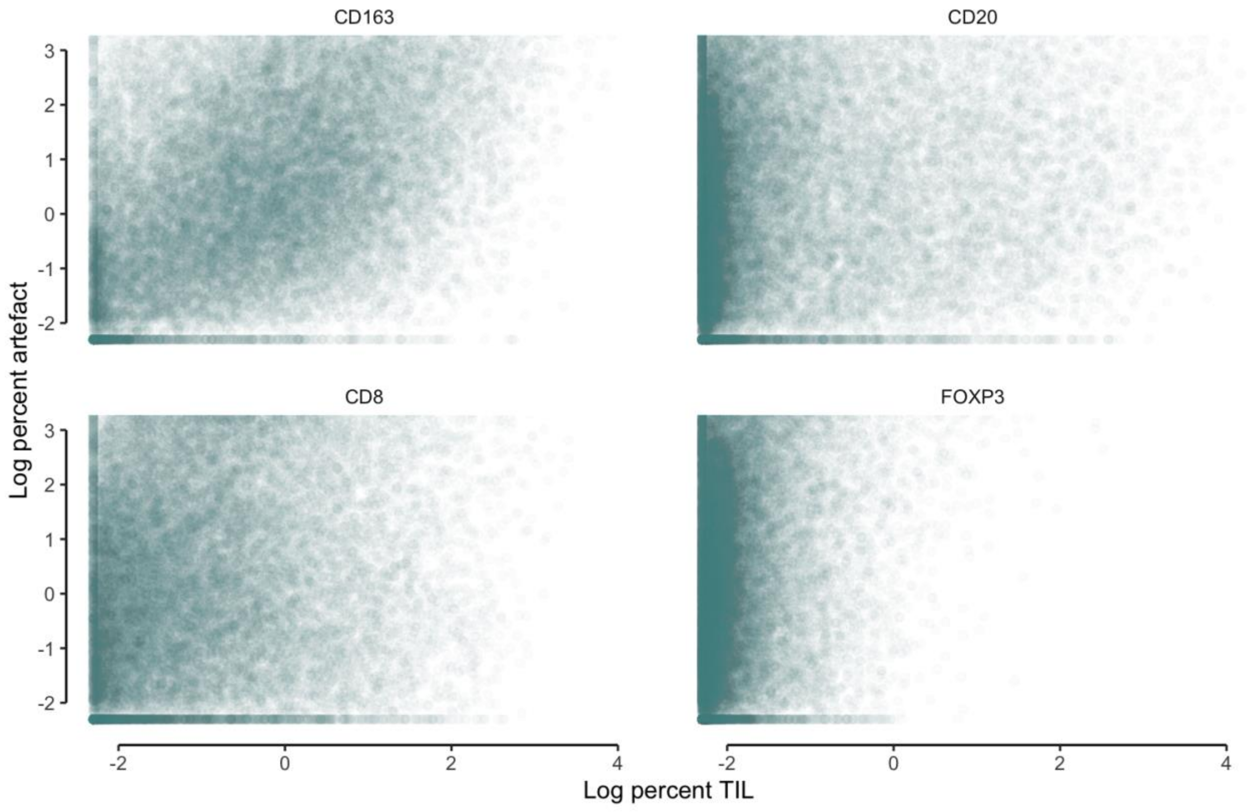

Supplementary figure 8: Scatterplot of total tissue area against percent TIIC by marker and tumour core size

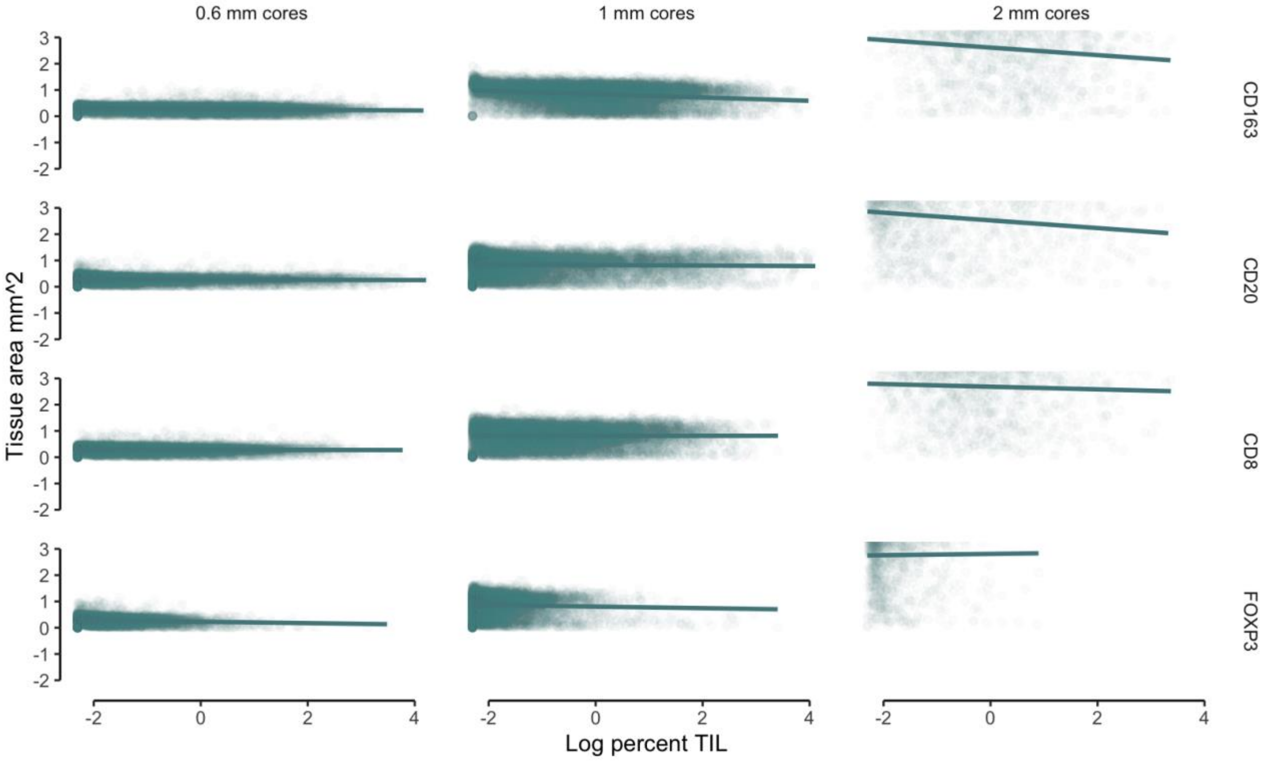


#### 4 Statistical methods

Schoenfeld residual tests were conducted on Cox regressions models with age, grade, tumour diameter, number of positive nodes and stratified by study (the set of variables used in the fully adjusted models), to test for violations of the proportional hazards assumption. The proportional hgazards assumption was violated (p<0.05) for grade in the ER-positive model and age, tumour diameter and grade for the ER-negative model. However, examination of the scaled residuals plotted against follow-up time revealed that the violation were only meaningful for grade in both ER-negative and ER-positive models (Supplementary figure 9 and Supplementary figure 10). We therefore fit an additional term for grade to allow the log hazard ratio to vary as a function of log time. We also tested the TIIC score variables for violations in each of the single marker partially adjusted models. Of the 24 models for log percent area at a core tissue area threshold of 0.25 $mm^2$, two were significant at a nominal P<0.05 (Supplementary table 5) and visual inspection of the scaled residuals showed that the violations were not substantial.

Supplementary figure 9: Scatterplot of Scaled Schoenfeld residuals against time for grade, number of positive nodes and tumour size in ER-negative Cox regression model

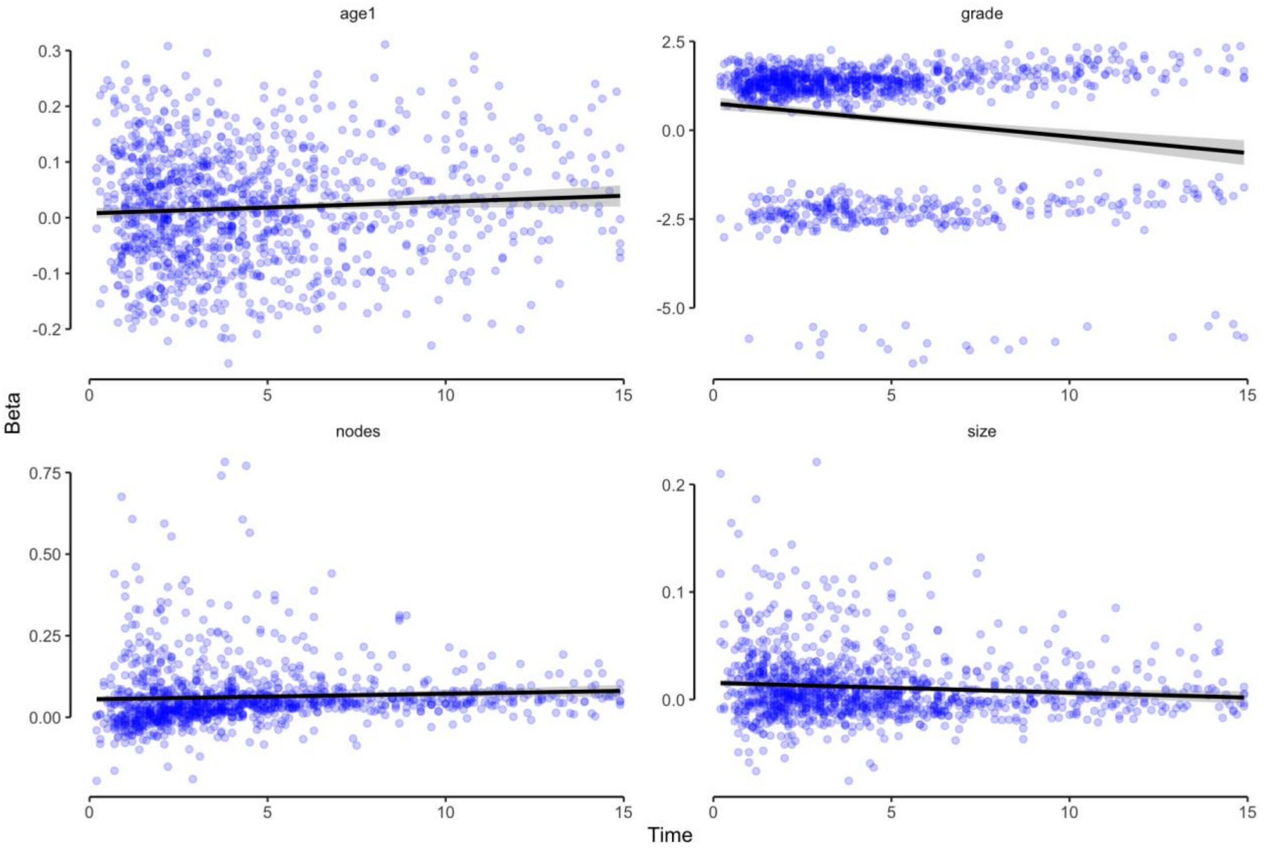


Supplementary figure 10: Scatterplot of Scaled Schoenfeld residuals against time for age at diagnosis, grade, number of positive nodes and tumour size in the ER-positive Cox regression model. age1 and age2 are the transformed variables for age at diagnosis (see main paper statistical methods)

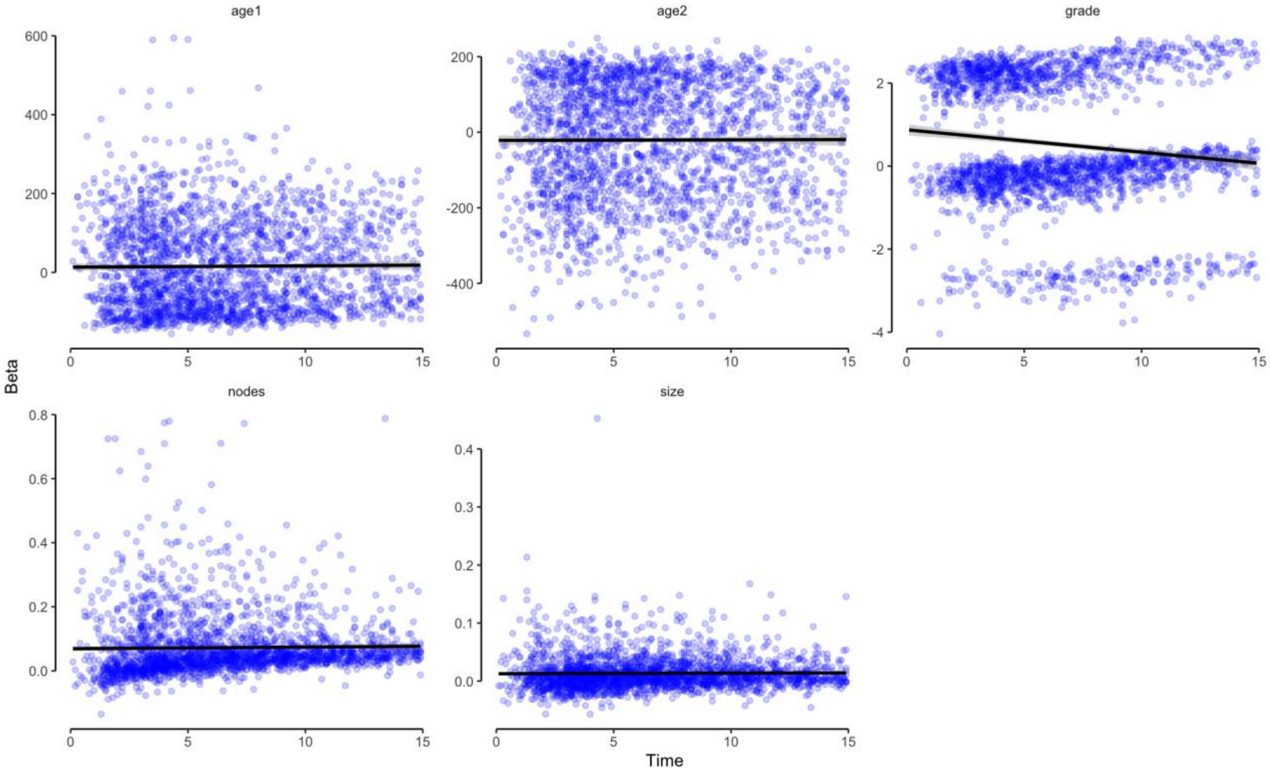
